# Implementing a Resource-Light and Low-Code Large Language Model System for Information Extraction from Mammography Reports: A Case Study

**DOI:** 10.1101/2025.04.08.25325371

**Authors:** Fabio Dennstädt, Simon Fauser, Nikola Cihoric, Max Schmerder, Paolo Lombardo, Grazia Maria Cereghetti, Sandro von Däniken, Thomas Minder, Jaro Meyer, Lawrence Chiang, Roberto Gaio, Luc Lerch, Irina Filchenko, Daniel Reichenpfader, Kerstin Denecke, Caslav Vojvodic, Igor Tatalovic, André Sander, Janna Hastings, Daniel M Aebersold, Hendrik von Tengg-Kobligk, Knud Nairz

## Abstract

**Background:** Large Language Models (LLMs) have been successfully used to extract structured data from free-text radiology reports. Most of current studies were conducted with private models accessed via Application Programming Interface (API). We aimed to evaluate the feasibility of using open-source LLMs, deployed on limited local hardware resources for extraction of structured information from free-text mammography reports, according to a Common Data Elements (CDE)-based framework.

**Methods:** Seventy-nine CDEs were defined by an interdisciplinary expert panel, reflecting real-world reporting practice. Sixty-one reports were classified by two independent researchers with 1533 classifications assigned to establish ground truth. Five different open-source LLMs deployable on a single GPU were used for data extraction using the *general-classifier* Python package. Extractions were performed for two different prompt approaches with classification metrics calculated overall and on subgroups. Additional analyses were conducted using thresholds for the relative probability of classifications.

**Results:** High inter-rater agreement was observed between manual classifiers (Cohen’s Kappa 0.83). Using default prompts, the LLMs achieved accuracies of 59.23–72.86%. Adapting prompts to better explain classification tasks improved performance for all models, with accuracies of 64.71–85.32%. Setting certainty thresholds further improved accuracies to >90% but reduced the coverage rate to <50%.

**Conclusion:** Locally deployed open-source LLMs can effectively extract information from mammography reports with good accuracy, addressing data privacy concerns while maintaining compatibility with limited computational resources. Prompt engineering substantially increases performance, highlighting the importance of optimization in clinical applications. Using a CDE-based framework provides clear semantics and structure, facilitating interoperability and consistent data extraction.

## INTRODUCTION

### Synoptic reporting and automated extraction of data from radiological documents

Synoptic or structured reporting is an approach to give a structured overview of a radiological report for a given radiological examination. Synoptic reporting has numerous advantages over free-text reporting, including improved consistency, clarity, and completeness of information [1]. Despite these advantages, synoptic reporting is not widely adopted in radiology [2] due to the increased time requirements, technological challenges, as well as the lack of standardization [3], [4].

Automated extraction of information from radiological reports is therefore of longstanding interest, as it could automatically generate corresponding synoptic reports in a standardized and precise way from free-text radiological documents [5]. Classical Natural Language Processing (NLP) techniques for text classification, like bag-of-words models or term-frequency-inversed document frequency (TF-IDF) have achieved success in extracting standardized data (e.g., Names, Dates, etc.) [6]. However, for more complex relevant information in radiological reports (e.g., peculiarities regarding certain morphological aspects etc.), the performance of such classical NLP approaches has been limited [7]. (Small) language models such as BERT, which, when given training for specific extraction tasks, can also perform very well, albeit not in a general muti-purpose instruction-following way [8] [9].

Recent advances in large language models (LLMs) opened new opportunities for automation of tasks that traditionally require human-level context understanding and reasoning. LLMs have been shown to be useful in structuring radiology reports [10], as well as in other settings, including writing clinical letters [11], clinical decision-support [12], medical education [13] or screening medical literature [14].

Furthermore, LLMs were shown to be highly effective for extracting information from unstructured clinical documents, substantially facilitating the automated generation of synoptic reports [15]. Several studies have demonstrated successful solutions using proprietary LLMs provided by private companies, such as OpenAI’s GPT-4 [16] or Anthropic’s Claude 3.5 [17], which are accessible via application programming interfaces (APIs). While such models demonstrate powerful performance, they rely on external data transmission thus raising critical concerns about clinical applications, particularly when dealing with sensitive patient data. To comply with data privacy regulations and prevent the transmission of sensitive information to external stakeholders, LLMs used within healthcare institutions should ideally operate on local hardware [18].

Therefore, the research for practical implementation is now focusing on locally deployed open-source LLMs for extraction of data from clinical documents [19]. However, many of the most capable LLMs are very large and for several tasks a correlation between model size and performance has been reported [20]. Many powerful models thus demand substantial computational resources and advanced hardware infrastructure to operate efficiently [21]. This highlights the importance of balancing technical feasibility with practical considerations in clinical applications [22].

### Clear data definitions using the Common Data Elements (CDE) concept

Regardless of the NLP technique used for data extraction, clearly defined data concepts with precise meanings are crucial. The National Institutes of Health (NIH) introduced the concept of Common Data Elements (CDEs) in 2011. As stated in the definition of the NIH a “*Common Data Element (CDE) is a standardized, precisely defined question, paired with a set of allowable responses, used systematically across different sites, studies, or clinical trials to ensure consistent data collection*” [23]. Initially designed for clinical trials, CDEs are increasingly applied for the collection of real-world data (RWD) [24].

Moreover, CDEs facilitate seamless implementation in information technology systems through standardized data formats such as JSON or XML, making them particularly relevant in the era of big data and machine learning [23].

For these reasons, CDEs were chosen as the fundamental semantic concept for collection and management of RWD within the SMARAGD (Smart Radiology Goes Digital) initiative [25]. SMARAGD is a collaborative effort between academia and industry, that leverages state-of-the-art NLP techniques to extract and manage clinical data, with a particular focus on the care pathway of breast cancer patients undergoing radiological examinations.

### Practical implementation of LLMs for automated information retrieval from mammography reports in a local setting

As part of the SMARAGD initiative, this case study focuses on the practical realization of an LLM-based system for extracting information from free text mammography reports. By using the modular and interoperable framework provided by CDEs, this system has been integrated into a broader data management architecture, demonstrating the potential of advanced NLP techniques in enhancing clinical workflows.

The primary objective of this work was to create a practical, easy-to-deploy, and cost-effective system for extracting categorical CDE values from free text radiology reports. This system ensures that all data is retained within the local environment and can be deployed on local hardware with limited computational resources. By leveraging open-source LLMs, the system eliminates external dependencies, effectively addressing data privacy concerns while also meeting the need for affordability in clinical applications.

## METHODS

### Development of CDE resources and data structure

As the first step, an interdisciplinary expert panel comprising physicians, healthcare informaticians, and computer scientists developed resources for generating synoptic reports with categorical (“Value List”) CDEs suitable for the clinical environment based on published NIH guidelines [23] [26].

The focus of CDE development was to ensure alignment with real-world clinical documentation and established reporting standards, specifically the guidelines of the American College of Radiology (ACR) on mammography reporting [27]. Based on previous research for defining common data elements of real-world clinical documents [28], the panel created and iteratively refined CDE resources until consensus was reached among the involved experts.

As the second step, CDEs were further evaluated by radiologists who used them in clinical practice for local documentation. Their feedback informed iterative revisions and the finalization of the data concepts. As the third step, data elements were organized into a hierarchical structure [29], consistent with the NIH’s CDE framework (corresponding to CDE forms [30]).

### LLM-based data extraction system in a local setting

#### General approach

As previously shown, LLMs can be used for effective data extraction from clinical documents [31]. For our use-case, the LLM was implemented as a basic data extraction and classification system as part of a broader framework, based on previous studies [14], [32]. We used the recently published *general-classifier* python package [33], which allows for easy-to-use deployment of LLMs for text classification. It facilitates the classification of a text (i.e., in our case the mammography report) into user-defined topics with mutually exclusive categories.

The Python package has been used for biomedical literature classification with high levels of accuracy with details on the functionality reported previously [32]. In brief, the library provides an easy-to-use interface for defining structured templates within which an LLM is executed; for each template, the LLM is provided with a prompt containing an instruction and the text from the medical document and is asked to answer a question about this text. This structure can be saved as a machine processable JSON file and later be imported for performing a classification. In general, LLMs function by predicting how a text (being a sequence minimal text-segments, so-called tokens) is likely to be continued. The probability of the next token to a given token sequence is calculated using a neural network and is based on the data used during model training. The probability of the next tokens associated to the value of given categories for a classification task can be calculated. The LLM can be used in this way as a general classification system by selecting the category with the highest probability.

Figure 1a illustrates the approach for a “Value List” CDE with three possible values (categories) out of which the LLM has to select the correct value. For each of the values the probability is calculated and the value with the highest relative probability is selected. An example for such a classification task might be the CDE “laterality of known breast cancer” with the possible values/answers “left”, “right”, and “unknown (no information provided in the document)”.

**Figure 1.**
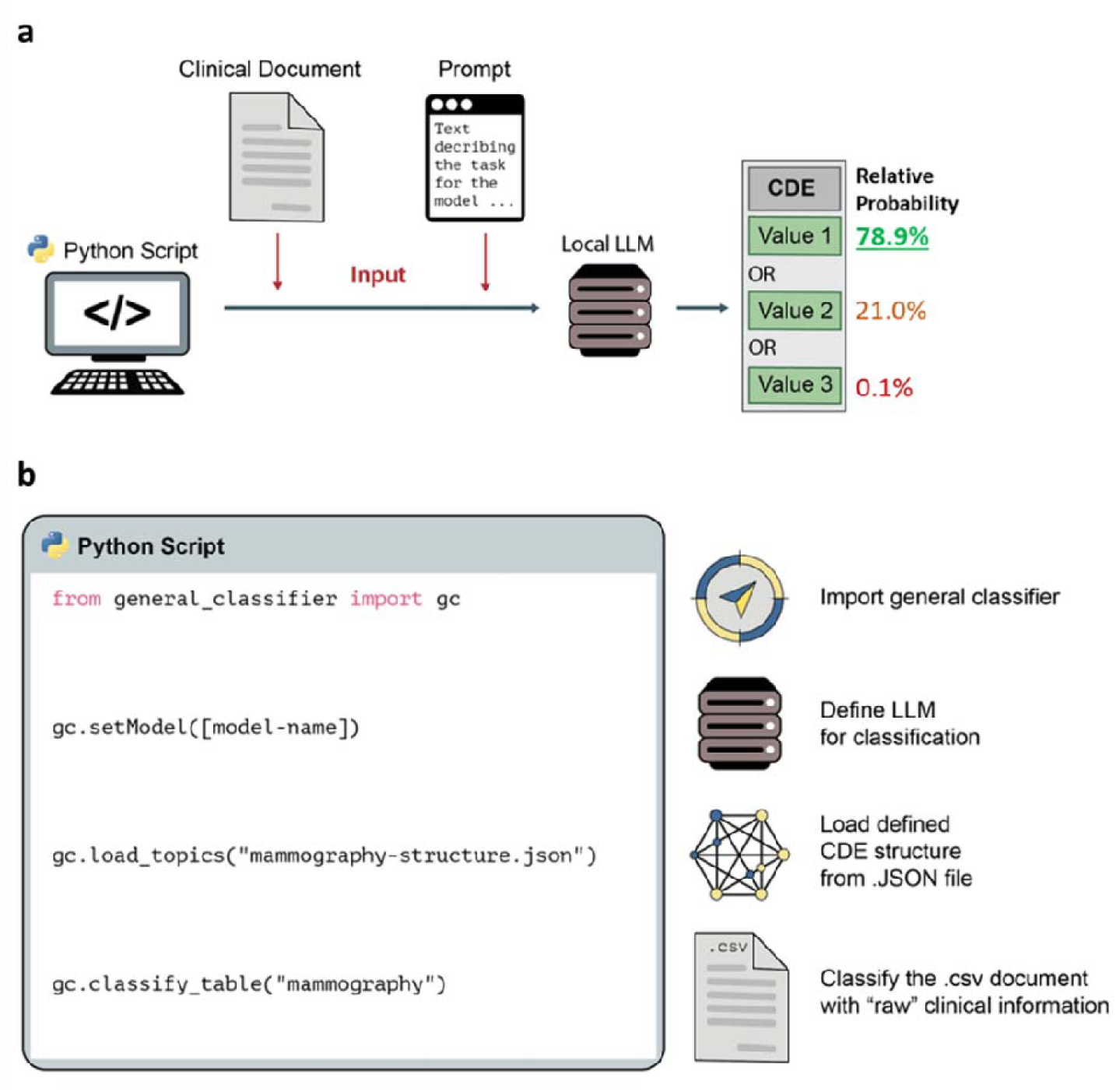
**a** Schematic illustration of the LLM-based approach for extracting the value of a categorical “Value List” CDE from a clinical document using LLMs. b – Principle of using the *general-classifier* framework for the data extraction. With three lines of code an LLM is initialized, the data structure (in .JSON) is loaded and the clinical documents (in .CSV) are analyzed accordingly.

#### Script and Hardware

A simple Python script was developed to implement the approach (Figure 1b; available on GitHub [34]). Based on the data structure for the reports a corresponding .JSON structure containing the classification topics, categories and hierarchies was created for the use of the *general-classifier* Python package (version 0.1.10).

The system was deployed on a local Linux server with an RTX6000 Nvidia GPU (with 48GB RAM) to ensure optimal processing capacity for running an open-source LLM. The software environment comprised Python (version 3.11.5) and PyTorch (version 2.4.0) [35] alongside dependencies specific to the open-source LLMs used.

#### Models

Five different modern open-source LLMs were used in the study. The LLMs were selected because of reported high performances on different relevant benchmarks such as MMLU [36], IFEval [37] and GPQA [38].

- Rombos-LLM-V2.6-Qwen-14b (Rombos) is a fine-tuned version of Alibaba’s Qwen2.5-14B model for precise instruction following [39]. The Qwen series excels in language understanding, generation, multilingual tasks, coding, math, and reasoning [35]. It has 14.8 billion parameters and a size of 29.57 GB.
- SOLAR Pro Preview Instruct (Solar Preview) is an instruction-tuned LLM by Upstage, reported to be the *“most intelligent LLM on a single GPU”* [40]. It’s a preview version of the upcoming SOLAR Pro model, optimized for following user instructions and generating helpful responses. It has 22.1 billion parameters and a size of 44.43 GB.
- Phi-4 is a compact AI language model developed by Microsoft released in February 2025 [41]. It delivers strong performance on reasoning, coding, and math tasks despite its small size. Trained on high-quality data, it offers capabilities comparable to much larger models while requiring fewer computational resources. It has 14.7 billion parameters and a size of 29.34 GB.
- Li-14B-v0.4 (Li) is an LLM developed by the Chinese company Century Innovation [42]. Like the Rombos model it is a fine-tuned version of the Qwen2.5-14B model. It strikes a balance between computational efficiency and performance capabilities. At the time of the study (state February 2025), it was ranked on the first place of LLMs up to 15B parameters on the Open LLM Leaderboard provided by Huggingface [43]. It has 14.8 billion parameters and a size of 29.52 GB.
- Lamarck 14B v0.7 (Lamarck) is a high-performing language model optimized for limited execution on hardware with limited resources [44]. Created through sophisticated merging techniques, it combines influences from multiple top models including Virtuoso-Small [45], DeepSeek [46], and DRT-o1 [47]. The model excels in multi-step reasoning, prose generation, and multilingual capabilities, using a custom toolchain of Low-Rank Adaptations (LoRAs) and targeted layer merges to achieve its balanced performance profile. It has 14.8 billion parameters and a size of 29.51 GB.

#### Prompting

As the LLM-based data extraction system depends on the output of the LLM, it also depends on the input prompt given to the model. As shown, the performance of LLMs on various tasks can be substantially increased by optimization of the prompt, so-called prompt engineering [48] [49]. In a first evaluation the default prompt for classification tasks provided in the *general-classifier* package was used. This default prompt contains a general instruction to conduct a classification with the name of the topic and the possible categories; see also [33]. For a second evaluation run, the prompts were adapted to better describe the classification task for individual CDEs.

The adapted prompts were systematically created mimicking a question-answer dialog using the following structure:

- INSTRUCTION – containing information that the task is to answer a specific question based on the information provided in a provided German mammography report
- QUESTION – containing the defined question that is to be answered.
- TEXT – the text of the mammography report; based on the *general-classifier* Python package the string “[TEXT]” is inserted, which will be replaced with the given text for the classification.
- ANSWER – starting the text answer which would be given to the question. This text ends the prompt and provides a direct context for the classification.

More sophisticated prompt-techniques (e.g., Chain-of-thought prompting or few-shot prompting) were not applied. The .JSON files containing all the default and adapted prompts used in the study are available on GitHub [34].

### Dataset and manual value assignment

For creation of the dataset used in the study, 61 anonymized mammography reports were used. Patients were referred to breast diagnostics concerning potential tumor lesions. Based on the resulting mammography reports, the task of obtaining the corresponding CDE values was assigned to two physicians independently. In case of disagreement between the two clinicians a final decision was made by the study coordinators. A final ground truth dataset with a total of 1533 CDE values assigned to the 61 reports was created. Conduction of the study was approved by the local ethics committee (BASEC Project ID 2022-01621).

### Statistical Analysis

The Inter-rater agreement (IRA) between the two researchers was calculated using Cohen’s Kappa [50]. The accuracy, as well as the micro-average values for recall and F1-Score were calculated for individual CDEs as well as for groups and subgroups defined in the data structure. It should be noted that not all CDEs had values for all reports in the dataset as some CDEs depended on the values of other CDEs within the hierarchy of the data structure (e.g. “laterality of previously conducted biopsy” is only provided if the CDE “previously conducted biopsy” is “true”).

The classification metrics were calculated on all evaluations given by the LLMs. In a second sub-analysis the accuracies were calculated only for the cases for which the LLM provided a high relative probability for the selected category (=probability for the selected category divided by the sum of probabilities for all categories). A threshold for this parameter which reflects the *“certainty of the model”* when performing the classification was set at 90%, 99% and 99.9% for additional sub-analyses. Apart from the classification accuracies, the proportion of classifications above the thresholds (coverage rate) was calculated.

### Time measurement and cost estimation of used hardware

The required time for the data extraction was calculated using the developed Python script [34]. A basic cost calculation of the used hardware infrastructure was conducted. A basic cost estimation for the used components was conducted in Swiss Francs, with conversions into euros and US dollars based on the exchange rates from January 16, 2025.

## RESULTS

### CDE-based data structure for the presentation of the mammography reports

The finalized CDE-based data structure consisted of 79 CDEs organized into two main groups and five sub-groups. The structure of the analyzed mammography reports included a pre-text (anamnesis) that outlines the patient’s history and reason for admission, followed by the main report text. Consequently, the two main groups of the data structure were defined as “Anamnesis,” with the sub-groups “Family anamnesis” and “Therapeutic anamnesis,” and “Report,” with the sub-groups “Breast composition,” “BIRADS,” and “Findings.” A schematic overview of the data structure with a translated artificial sample of a mammography report is provided in Figure 2.

**Figure 2:**
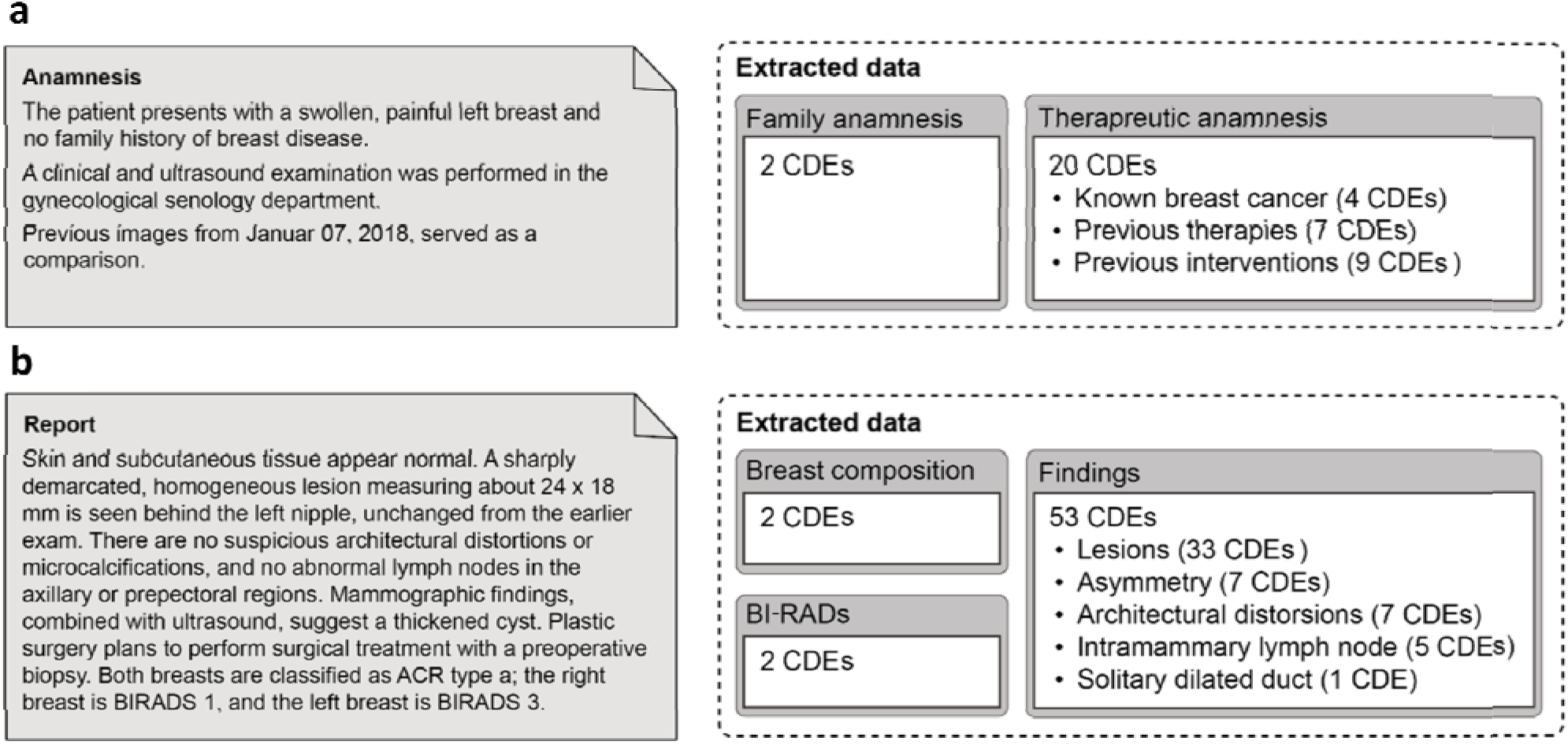
Basic hierarchical structure for the data extracted from the mammography reports. Grouping of CDEs with the number of CDEs per group and subgroup.

A detailed list of CDEs and data structure is provided in **Appendix 1. Interrater agreement** A high level of IRA with a Cohen’s Kappa of 0.83 was observed between the two researchers doing the manual value assignment. The smallest level of agreement was seen for the group “Findings” with a value of 0.71. An overview of the values for the individual groups is provided in **Table 1**. Detailed results on all individual CDEs are provided in **Supplementary Table 1**.

**Table 1:**
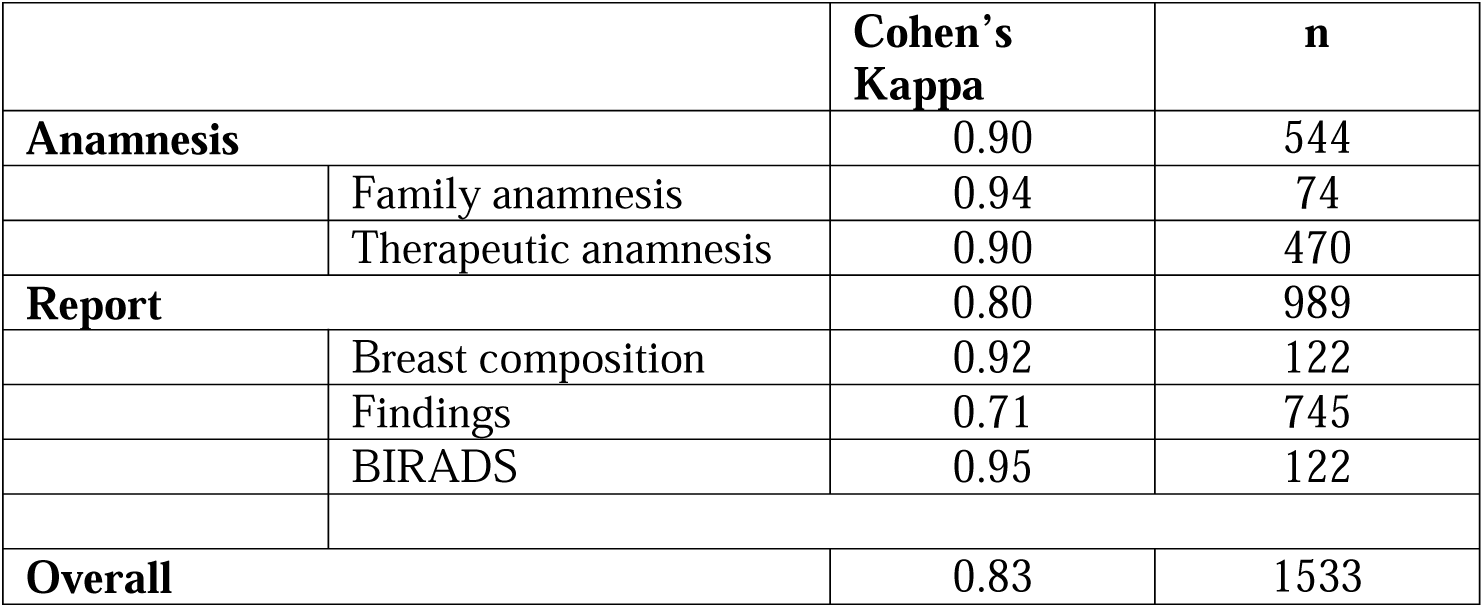
Inter-rater agreement on the different (sub-)groups of CDEs between the two independent researchers assigning the values.

### Performance of the LLM-based data extraction with default prompts

Using the same default classification prompt of the *general-classifier* Python package (See also [33]) for all CDEs, the system achieved an overall accuracy of 72.86% for Rombos, of 61.58% for Solar Preview, of 59.23 % for Phi-4, of 64.71% for Li and of 68.49% for Lamarck. Further details with results on the individual groups and subgroups are presented in Figure 3. The results regarding Micro Recall and Micro F1-Score are provided in the **Supplementary** Figures 1-2.

**Figure 3:**
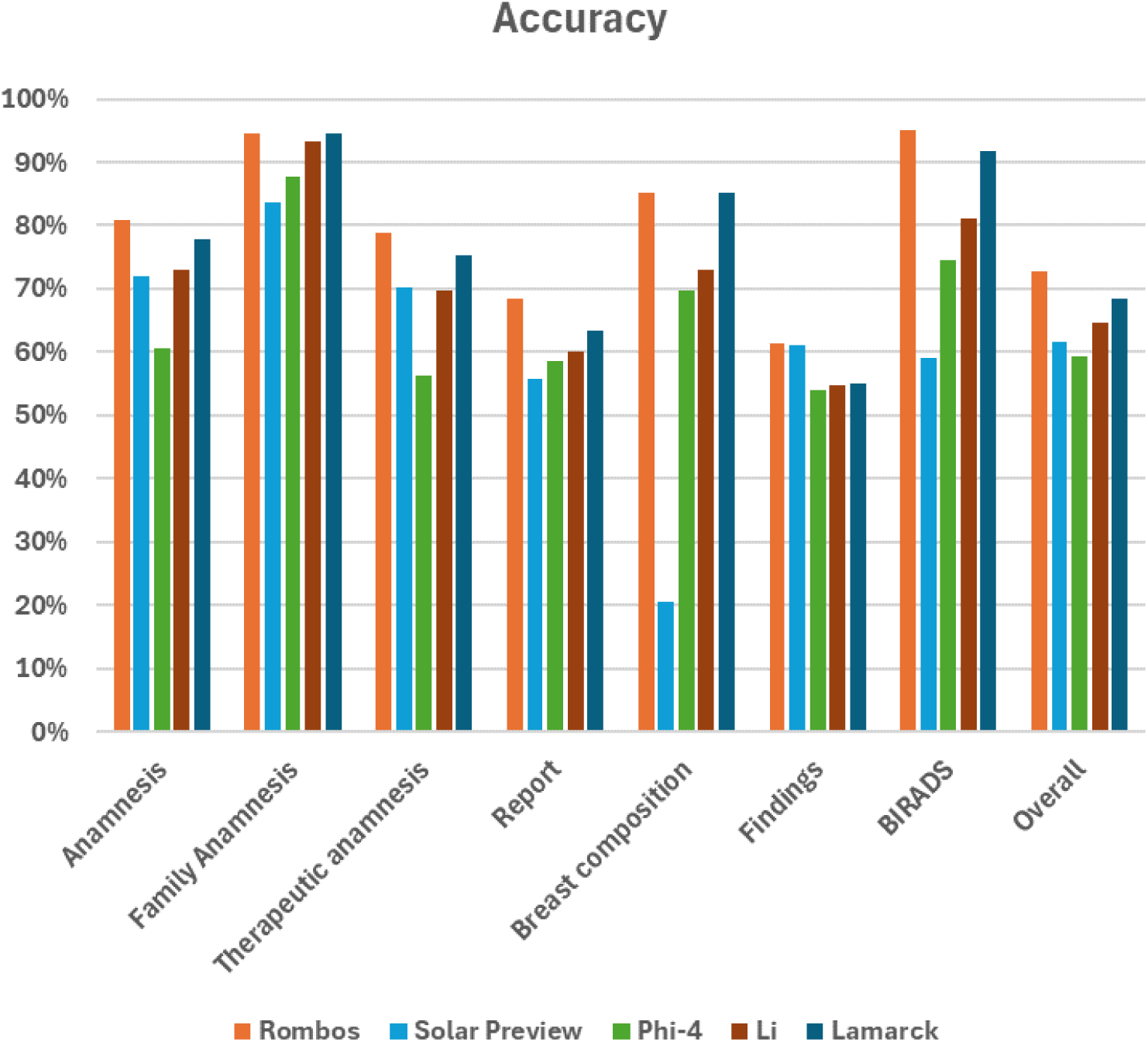
Accuracy of the LLM-based classification system with the five LLMs on the different groups, sub-groups and overall in classifying the mammography reports. Results for the default prompt.

### Performance of the LLM-based data extraction with adapted prompts

Adaptation of the prompts to better explain the classification tasks improved the performance of th system for all models. A comparison between default and adapted prompt for the CDE “Previousl conducted mammography mentioned?” is provided in Figure 4, for which accuracies increase from 55.74 – 72.13% to 62.30 – 96.72%.

**Figure 4:**
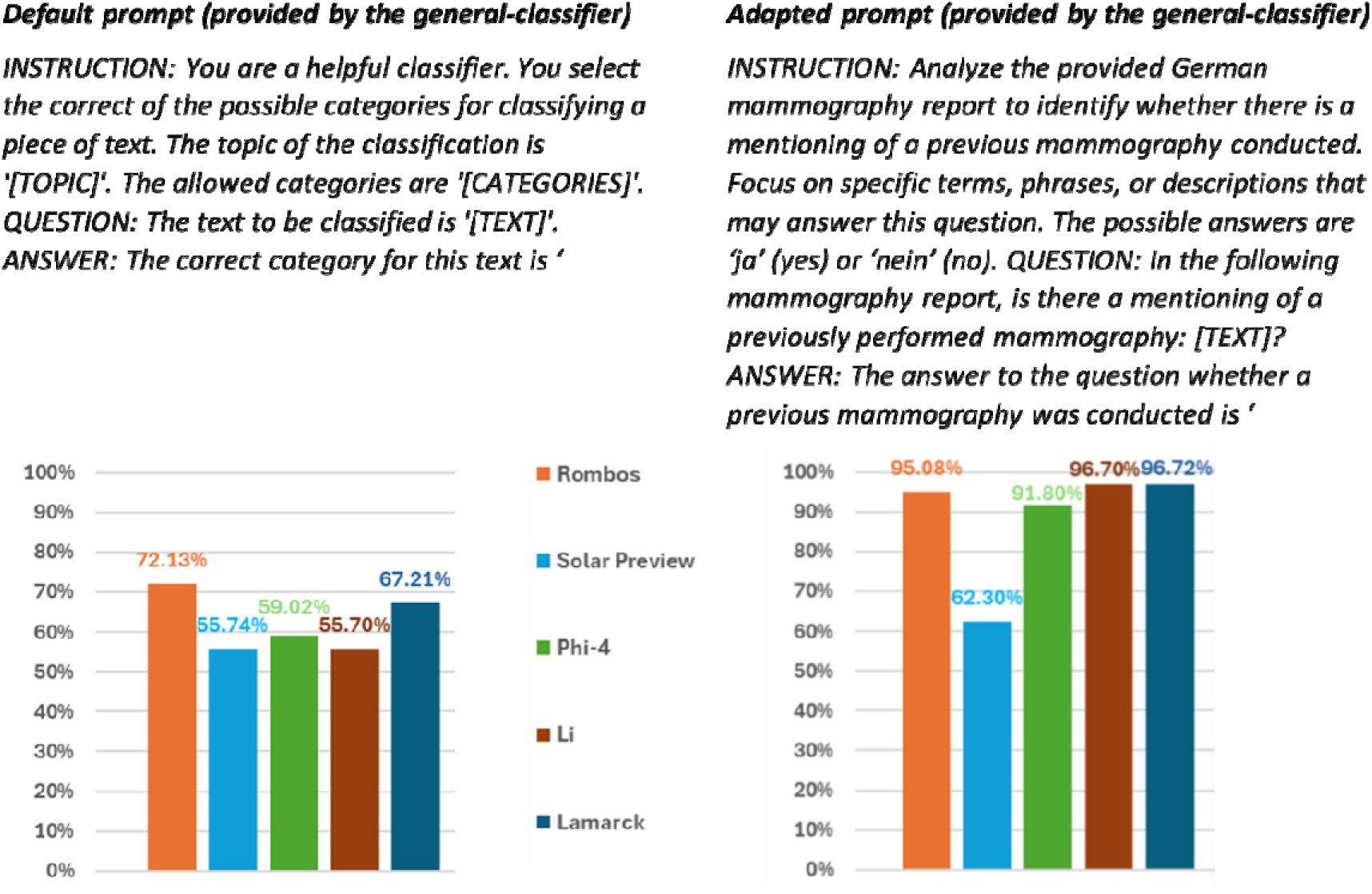
Comparison of default prompt provided by the general-classifier and adapted prompt better explaining the classification task for the CDE “Previously conducted mammography mentioned?”. Comparisons of accuracies for the different LLMs. The general-classifier replaces [TOPIC] with the classification topic „Zuvor erfolgte Mammographie erwähnt?“ (=”Previously conducted mammography mentioned?”) and [CATEGORIES] with the classification categories „ja, nein” (=”yes, no”).

Using the adapted prompts, the system reached overall accuracies of 85.32% for Rombos, of 64.71% for Solar Preview, of 65.04% for Phi-4, of 77.43% for Li and of 81.60% for Lamarck. Detailed results are presented in the **Supplementary** Figures 3-5.

### Sub-analysis with threshold for relative probability

In the sub-analysis using a threshold for the relative probability (=*“certainty”*), an increase of the accuracy compared to the baseline was seen across all models and thresholds.

Using the default prompt, compared to baseline, focusing only on the cases with >99.9% relative probability led to 86.50% accuracy (+13.64 %; coverage rate of 43.97%) for Rombos, to 94.38% for Solar-Preview-Instruct (+29.04%; coverage rate of 10.44%), to 80.00% for phi-4 (+32.50%; coverage rate of 1.96%) and to 79.86% for Lamarck (+11.37%; coverage rate of 37.90%).

Similar improvements were observed when using the adapted prompts with accuracies of 94.53% (+9.21%; 43.97% coverage rate) for Rombos, of 93.75% for Solar Preview (+29.04%; coverage rate of 12.52%), of 97.54% for Phi-4 (+32.50%; coverage rate of 7.96%), of 92.43% for Li (+15.00%; coverage rate of 20.68%) and of 91.72% for Lamarck (+10.12%; coverage rate of 39.40%). The results are presented in Figure 5 (default prompt) and Figure 6 (adapted prompt) for Rombos as well as in the **Supplementary** Figures 6-13 for the other LLMs.

**Figure 5:**
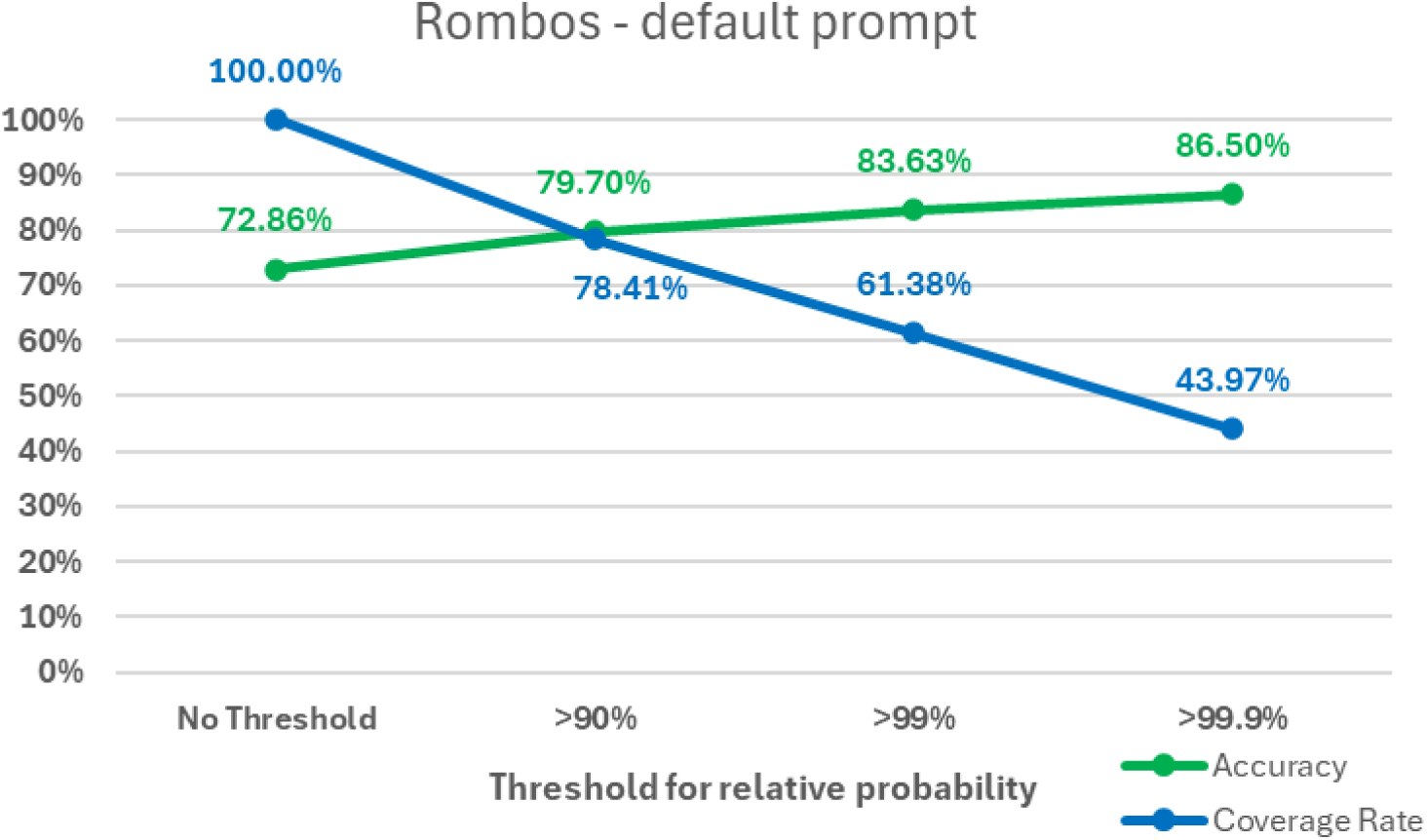
Rate of accuracy and coverage for the classifications using the default prompt with the Rombos model, depending on the threshold for the relative probability.

**Figure 6:**
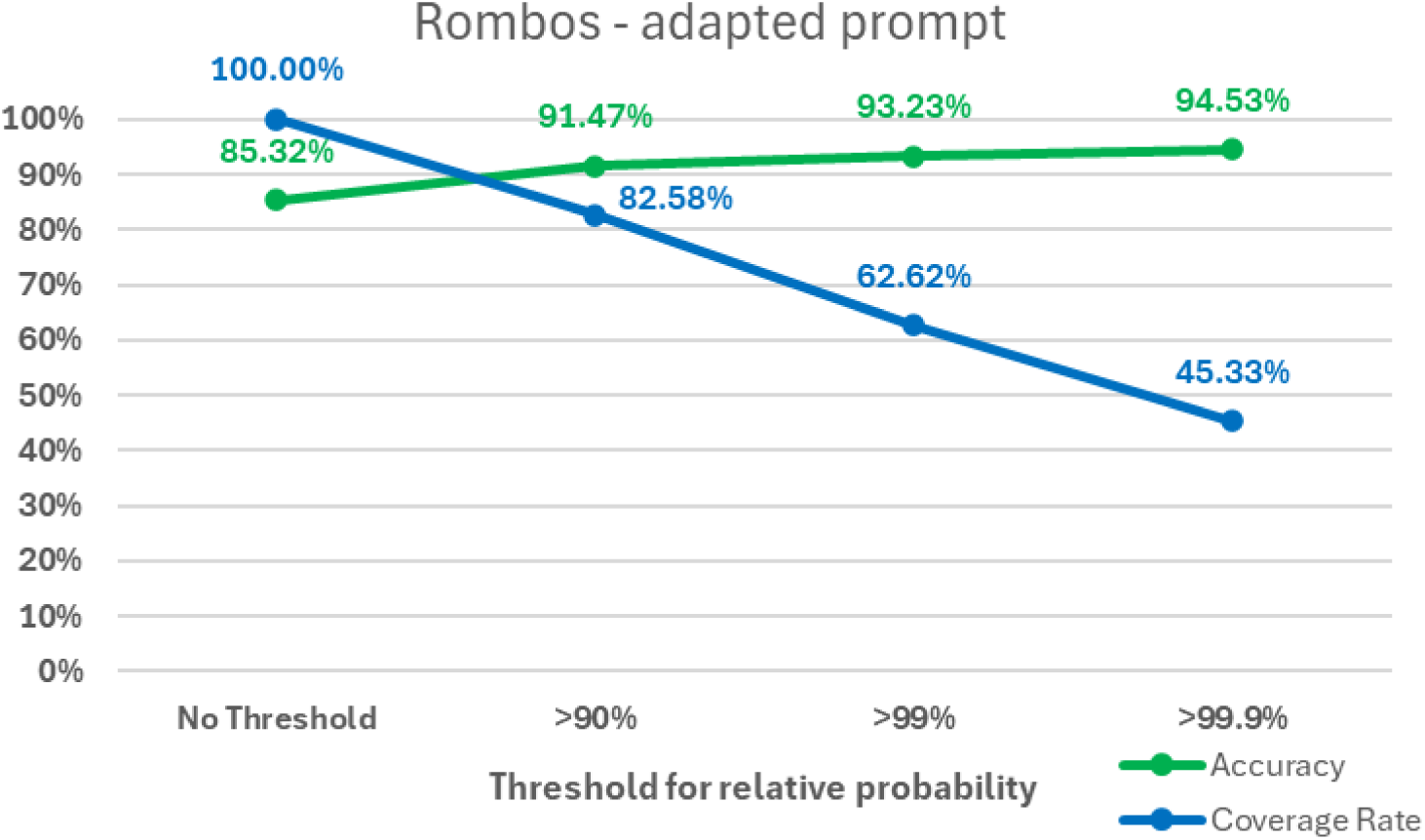
Rate of accuracy and coverage for the classifications using the adapted prompt with the Rombos model, depending on the threshold for the relative probability.

### Time and costs

For the default prompt, the script was executed in 2665.69 seconds for Rombos (43.70 seconds/report), in 6581.68 seconds for Solar-Preview (107.90 seconds/report), in 2489.52 seconds for phi-4 (40.81 seconds/report), in 2625.25 seconds for Li (43.04 seconds/report) and in 2746.58 seconds for Lamarck (45.03 seconds/report).

Similar results were obtained for the adapted prompts with 2614.33 seconds for Rombos (42.86 seconds/report), 6491.45 seconds for Solar-Preview (106.42 seconds/report), 2423.45 seconds for phi-4 (48.08 seconds/report), 2607.59 seconds for Li (42.75 seconds/report) and 2718.58 seconds (39.73 seconds/report) for Lamarck. The overall hardware costs of the used components were estimated at 7600 CHF with details provided in **Table 2**.

**Table 2:** Basic cost estimation of the used hardware components.

|  | Costs in CHF |
| --- | --- |
| **Core Components** |  |
| CPU: Intel Core i7-13700K | ≈350 |
| Motherboard: Z790 | ≈250 |
| RAM: 64 GB DDR5 | ≈320 |
| Storage: 1 TB NVMe SSD | ≈120 |
| GPU: NVIDIA RTX 6000 (Ada)<br>With 48GB on-board RAM | ≈6000 |
| PSU: 1000W | ≈190 |
| Case / Chassis: | ≈100 |
| Cooling | ≈80 |
| **Peripherals** |  |
| Monitor | ≈150 |
| Keyboard & Mouse | ≈40 |
| **Total** | ≈7600 (ca. 8100 € or 8300 USD) |

## DISCUSSION

### LLM-based data extraction

Due to the recent advancements in the performance of LLMs these models can now successfully perform various clinical tasks that for a human would require reasoning and contextual understanding. Various groups have demonstrated successful extraction of clinical data from mammography, CT and MRI reports [31] [51] [52]. Many of these studies have used closed LLMs only accessible via API, such as OpenAI’s GPT models. While API-based models provide a ready-to-use and affordable opportunity to implement LLMs, such systems rely on external stakeholders and (at least anonymized) data has to be sent outside of the local data environment. To avoid external transmission of data the research interest has grown in using locally deployed LLMs for data extraction [53] [54]. In a recent study Woźnicki et al. used the open-source LLM Llama-2-70B-chat to extract data from chest radiograph reports [19]. They achieved an F1 score of 0.70 for English and 0.68 for German reports and, similarly to our study, noticed considerable variations in performance and understanding of semantics depending on the data to be extracted. Obviously, these results are not comparable, since different data fields were extracted and different datasets were used. However, the fact that we were able to achieve similar high accuracies and F1-scores with LLMs having [– ⅓ of the parameters, aligns with the overall trend in LLM research with models rapidly becoming more efficient and more powerful [55]. While the work of Woźnicki et al. was published just a few months ago, Llama-2-70B-chat is not considered anymore state-of-the-art with Llama-3 [56] and other open-source models like DeepSeek-R1 [46] released, demonstrating the rapid pace of development in the field.

As part of the SMARAGD initiative, we developed an LLM-based data extraction system using an open-source model executed on local hardware. We also focused on the practical implementation considering environments of limited hardware resources and addressing data privacy issues by keeping medical data within a local environment.

### Performance and practical implementation

Even without any optimization, an accuracy of >70% in answering medical questions about a report was achieved in our study, with the possibility to adapt the prompts and improve the performance. However, the system failed to consistently give the correct answers. Since a perfect system with 100% accuracy will not be possible, it is therefore important to mitigate the risks and better understand the situations when there is a high risk of incorrect classifications. The possibility to calculate the “relative probability” for a category, resembling the “certainty” of an LLM when doing a classification, is very valuable for this purpose. As we have seen, setting a threshold with high certainty of 90%, 99% or 99.9% can indeed be used to achieve higher performances. Depending on how critical a classification is, higher thresholds could be demanded. While this does not completely solve the issue (and may lead to an unacceptable low coverage rate), the risks can be mitigated. From a practical point of view, classifications with *“low certainty”* could be flagged for human verification.

Some of the lowest levels of performance were seen for the group “Findings”, which is also the group with the lowest IRA. This might indicate that questions that are less clear and difficult to answer for humans are ambiguous for LLMs as well.

It should be noted that the approach itself can be easily applied to many other documents and scenarios (Figure 1b). As we have seen, a performant LLM-based system for data extraction from free text of mammography reports can be realized with a budget for the hardware components of less than 8000 CHF. While we used an NVIDIA RTX 6000 (Ada) with 48GB on-board RAM, the Rombos, Phi-4, Li and Lamarck models could have also been executed on a smaller (and cheaper) GPU with 32 GB on-board RAM. Of course, this cost estimation only includes the hardware and no personnel costs.

The classification script itself is easy-to-deploy using only a few lines of code [34]. The source code for the project is publicly available under an open-source license and can be directly implemented for other scenarios of data extraction after adjustment of the relevant topics and categories.

We hope that this work serves as a guide for researchers and clinicians interested in setting up a local LLM-based data extraction system. Active and practical participation of clinicians in the implementation of AI into healthcare systems will be crucial to ensure the systems meet real-world requirements and ultimately lead to the best possible outcome for patients [57].

### CDEs for synoptic reporting and IT integration

The performance of the system depends on the individual question asked (or CDE defined). Having a clear data structure is fundamental for structuring medical information. It further helps to identify for which groups and CDEs the system performs well and for which it does not. CDEs are increasingly relevant for collecting medical data, not only within scientific trials, but also in clinical practice [24]. Particularly for RWD in radiology, CDEs are highly valuable, as they provide clear semantics and structure for documentation [58]. They are increasingly being used for integration of radiological AI systems [59]. Due to the increasing relevance of CDEs in radiology, the Radiological Society of North America (RSNA) in collaboration with the ACR have started developing their own repository of radiological CDEs [60]. CDEs can build a shared framework for synoptic reporting, similar as the datasets of the International Collaboration on Cancer Reporting (ICCR) are used for structured reporting in pathology [61] (in principle the ICCR datasets are equivalent to the CDE concept defined by the NIH).

Furthermore, as CDEs are directly machine-readable, they can be used for implementation of information technology systems (including generative AI) [62]. LLM-based as well as classical NLP systems for data extraction can directly be linked [63]. As reported, the CDE concept can be successfully applied for collection of RWD via implementation into clinical IT and AI systems [64], [28].

CDEs provide an ideal means to support data interoperability. Rather than seeing radiology reports as a separate entity, information from different medical disciplines can be converted into CDE values. Combining extracted CDE values from various diagnostic and therapeutic paths thus may be a solution to cope with the challenges of data-driven medicine [25].

### Limitations

Our study has several limitations. In comparison to other approaches for data extraction like rule-based systems or named-entity recognition (NER), LLMs are black boxes, and it is very challenging to understand exactly, why a conclusion was reached by the model. There might be ways address this using systems like the LLM Transparency Tool of Meta [65], but the issue cannot be completely solved. Furthermore, LLMs are way more resource intensive and some of the CDE values in our study could probably also have been accurately extracted using rule-based systems.

It should also be noted that the script used in this study can only extract categorical data values (”Value List CDEs”). To retrieve other relevant information (e.g., number values) further adaptation would be needed.

Furthermore, while 61 mammography reports from clinical practice were used, no external validation of the performance with a second independent dataset was done. While 1,533 classification tasks were performed by two independent researchers and the LLMs, the number of classifications for each individual CDE is limited, which restricts the ability to draw more comprehensive conclusion. Generalization of the results may therefore be limited. However, it is likely that the general approach would work well in other settings, as the used LLMs are general models and already perform well without specific optimization for our use-case. It should be noted that the LLMs used in the study were not specifically trained on internal data. The recent literature on LLM-based data extraction furthermore suggests that this kind of approach successfully works in various settings [31].

One strength of our study is the clear definition of data concepts within a structured hierarchy using the CDE concept. If clear data standards are used on a broader level this can be used to facilitate interoperability, clear semantics and comparison of results.

## CONCLUSIONS

Locally deployed, open-source LLMs can effectively extract information from unstructured mammography reports for structuring within a CDE-based framework. By retaining all data locally, the approach addresses key data privacy concerns and remains compatible with settings that have limited computational resources. Prompt engineering substantially increases accuracy and highlights the importance of iterative optimization in real-world clinical applications. Going forward, wider adoption of standardized data definitions and locally implementable LLMs holds promise for enhancing interoperability, streamlining clinical workflows, and advancing research through reliable and secure data extraction.

## List of abbreviations

ACR: American College of Radiology
AI: Artificial Intelligence
API: Application Programming Interface
CDE: Common Data Element
IRA: Inter-rater agreement
IT: information technology
LLM: Large Language Model
LoRA: Low-Rank Adaptation
NER: Named entity recognition
NIH: National Institutes of Health
NLP: Natural Language Processing
RSNA: Radiological Society of North America
RWD: Real-world Data

## Code and data availability statement

The source code for the project as well as the .JSON files containing the relevant data about the data structure with classification topics, categories and prompts are provided at *GitHub* [34]. Information regarding the used anonymized mammography reports can be obtained from the authors upon reasonable request.

## Author contributions

Conceptualization – FD, NC, MS, HVTK, DA, SD, DR, KD, KN, GM

Data structuring – FD, SF, NC, MS, HVTK, PL, SD, TM, JM, LC, RG, LL, HB, CV, IT, AS, DR, KD, KN

Methodology, technical implementation – FD, MS, JM, LC, RG, LL

Methodology, creation of datasets – SF, TM Statistical Analysis – FD

Writing, original draft preparation – FD, NC, MS, KN Writing, illustrations – FD, IF

Writing, review and editing – All authors Project administration – FD, NC, KN

## Data Availability

The source code for the project as well as the .JSON files containing the relevant data about the data structure with classification topics, categories and prompts are provided at GitHub (https://github.com/Smart-Radiology-Goes-Digital-SMARAGD/low-code-LLM-CDE-extraction-mammography). Information regarding the used anonymized mammography reports can be obtained from the authors upon reasonable request.

## Acknowledgement

Not applicable.

## Ethics approval

An ethics approval for the study was granted by the Ethics Committee of the Canton of Bern (BASEC number 2022-01621), in alignment with the principles outlined in the Declaration of Helsinki.

## Informed consent

The data utilized in this study comprised anonymized mammography reports. All patients provided informed consent for the use of their data for research purposes, in accordance with ethical standards.

