## Supplementary Figures for "Implementing a Resource-Light and Low-Code Large Language Model System for Information Extraction from Mammography Reports: A Case Study"

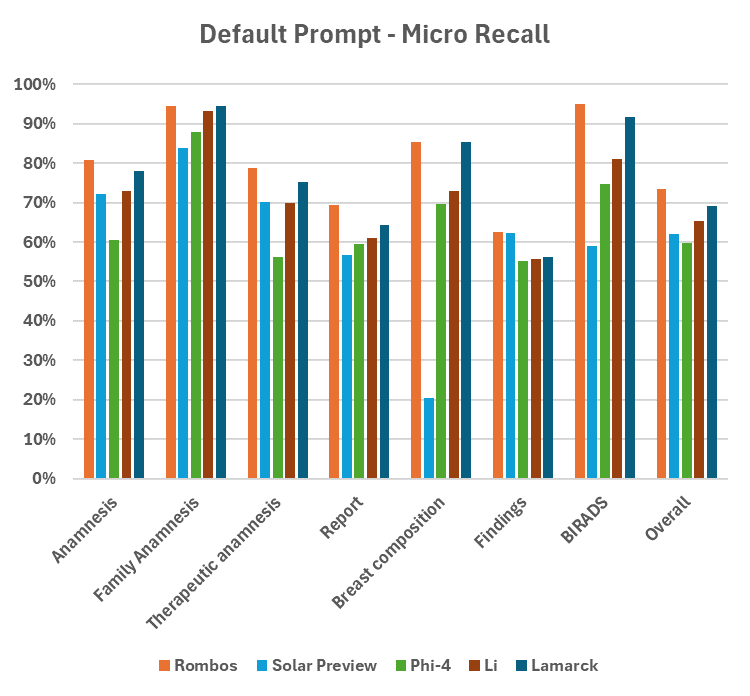

Supplementary Figure 1: Micro-Recall of the LLM-based classification system with the five LLMs on the different groups, sub-groups and overall in classifying the mammography reports. Results for the standard prompt.

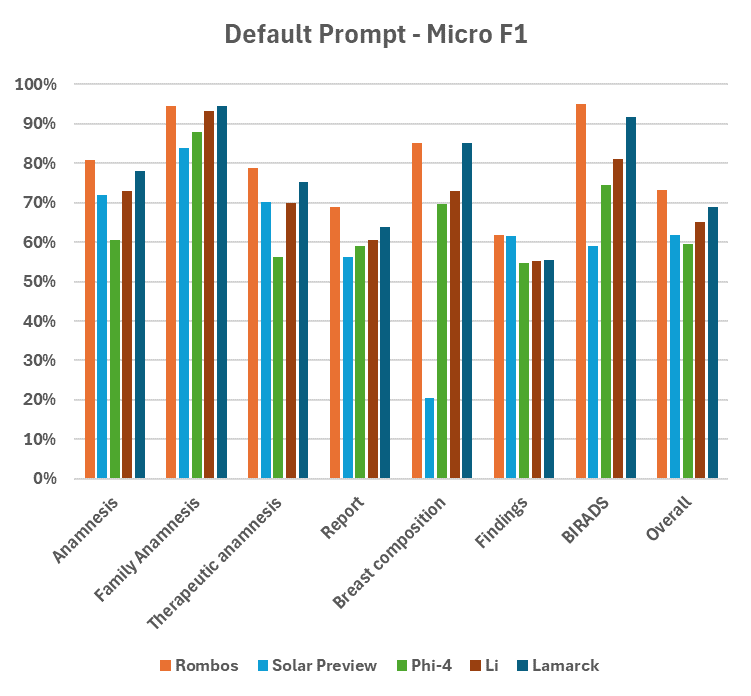

Supplementary Figure 2: Micro-F1 of the LLM-based classification system with the five LLMs on the different groups, sub-groups and overall in classifying the mammography reports. Results for the standard prompt.

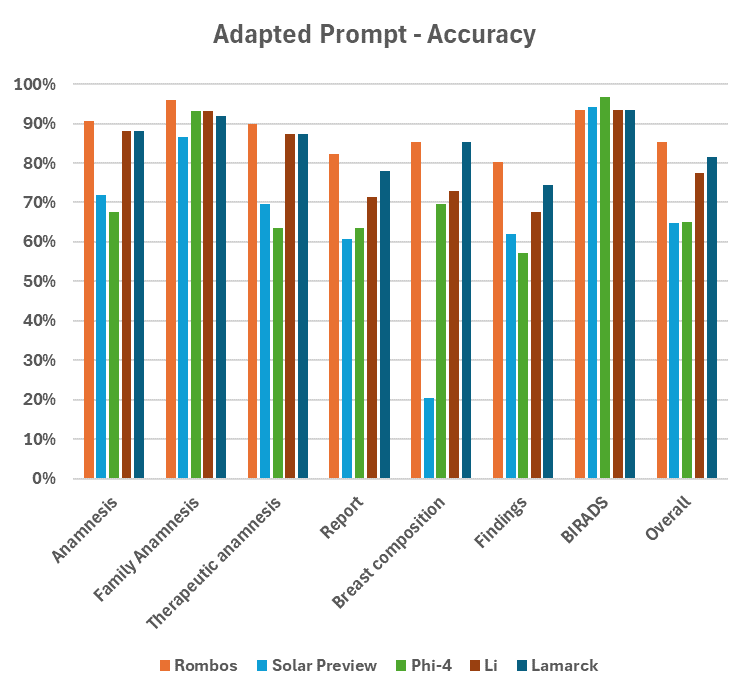

Supplementary Figure 3: Accuracy of the LLM-based classification system with the five LLMs on the different groups, sub-groups and overall in classifying the mammography reports. Results for the adapted prompt.

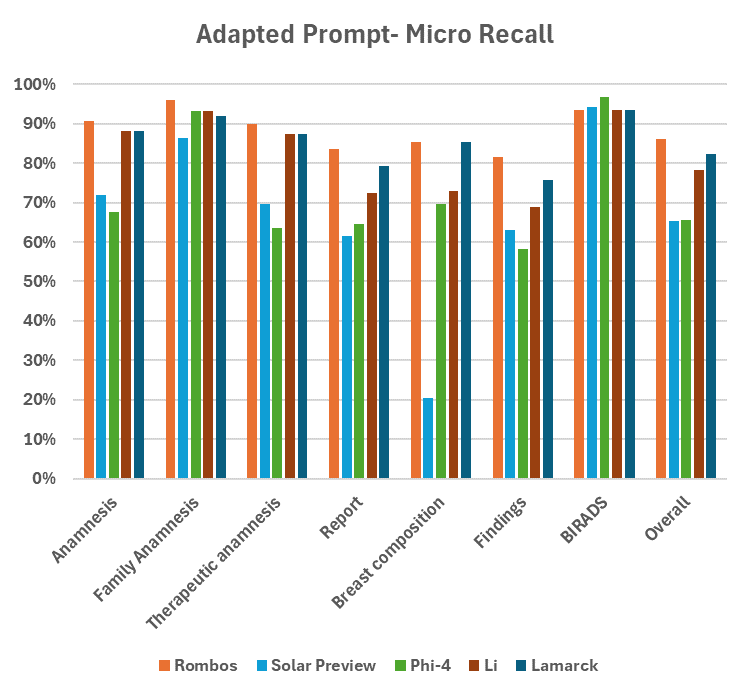

Supplementary Figure 4: Micro Recall of the LLM-based classification system with the five LLMs on the different groups, sub-groups and overall in classifying the mammography reports. Results for the adapted prompt.

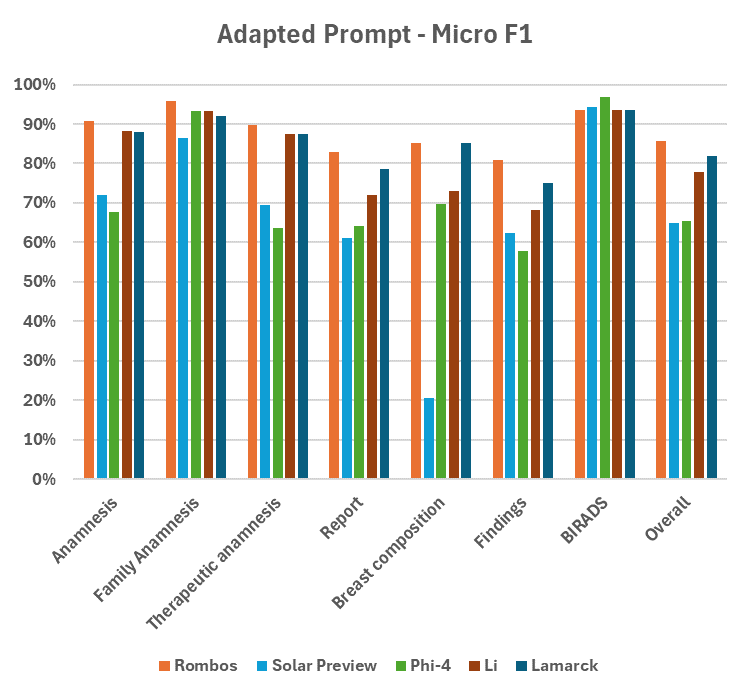

Supplementary Figure 5: Micro F1 of the LLM-based classification system with the five LLMs on the different groups, sub-groups and overall in classifying the mammography reports. Results for the adapted prompt.

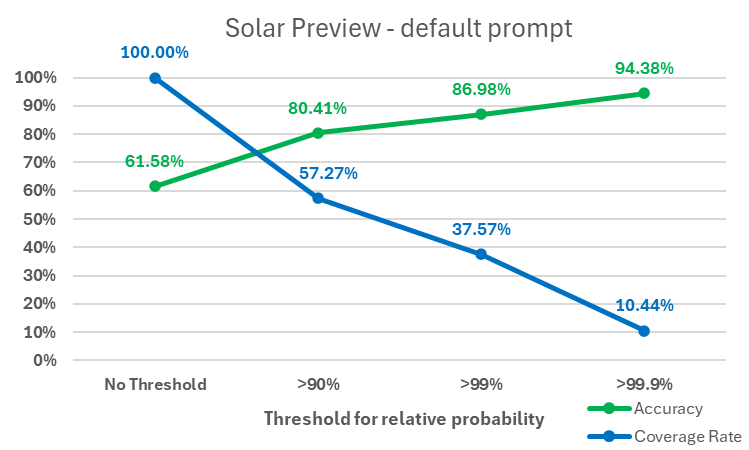

Supplementary Figure 6: Rate of accuracy and coverage for the classifications using the standard prompt with the Solar Preview model, depending on the threshold for the relative probability.

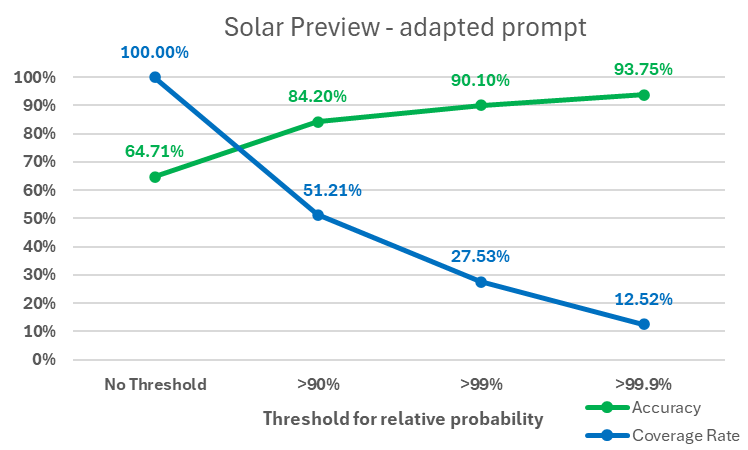

Supplementary Figure 7: Rate of accuracy and coverage for the classifications using the adapted prompt with the Solar Preview model, depending on the threshold for the relative probability.

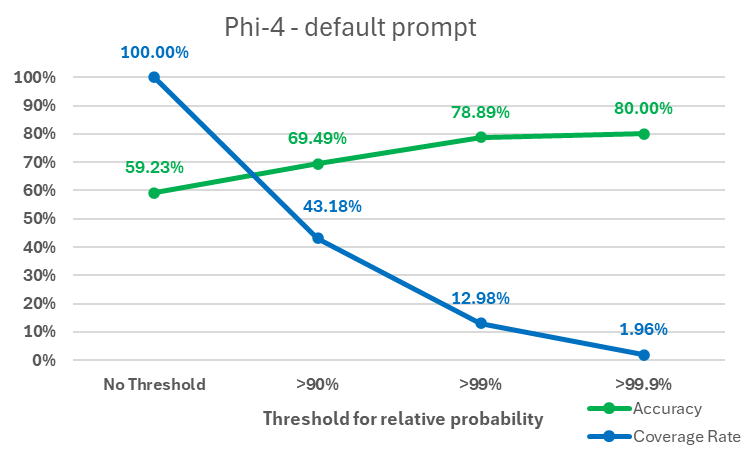

Supplementary Figure 8: Rate of accuracy and coverage for the classifications using the standard prompt with the phi-4 model, depending on the threshold for the relative probability.

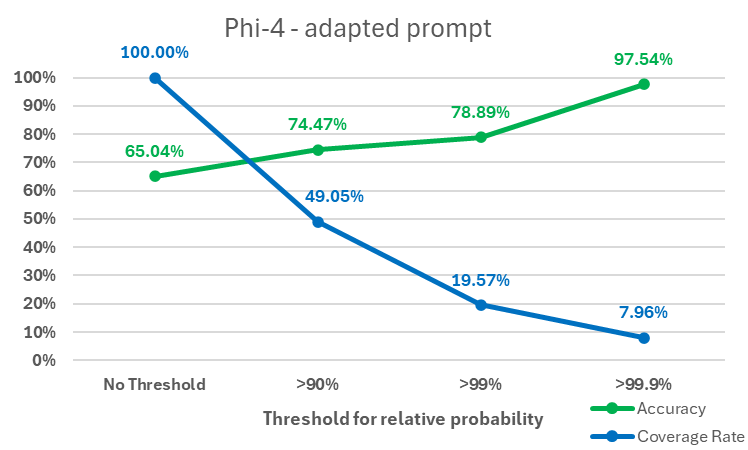

Supplementary Figure 9: Rate of accuracy and coverage for the classifications using the adapted prompt with the phi-4 model, depending on the threshold for the relative probability.

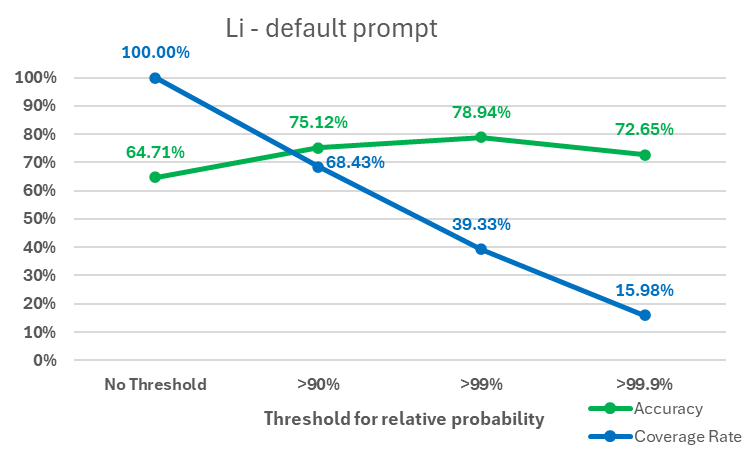

Supplementary Figure 10: Rate of accuracy and coverage for the classifications using the adapted prompt with the Li-14B model, depending on the threshold for the relative probability.

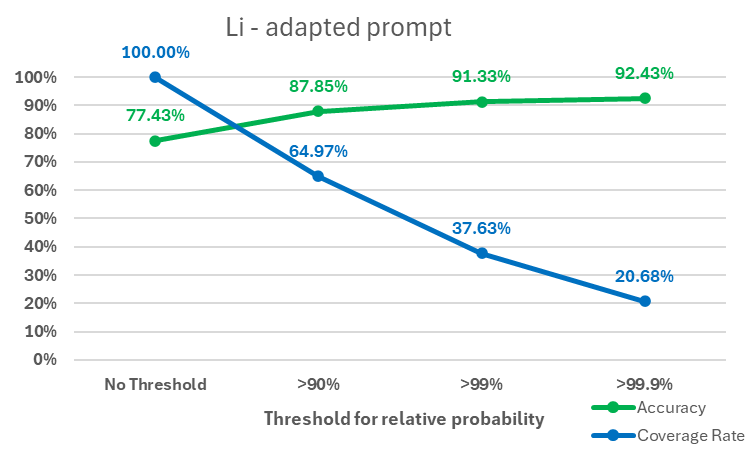

Supplementary Figure 11: Rate of accuracy and coverage for the classifications using the adapted prompt with the Li-14B model, depending on the threshold for the relative probability.

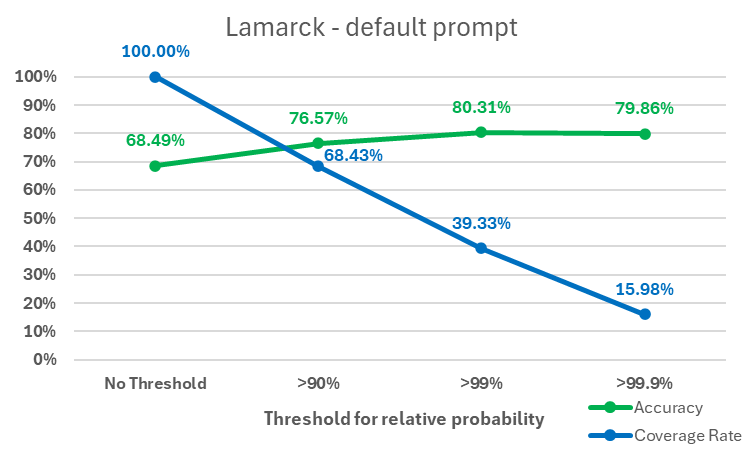

Supplementary Figure 12: Rate of accuracy and coverage for the classifications using the adapted prompt with the Lamarck model, depending on the threshold for the relative probability.

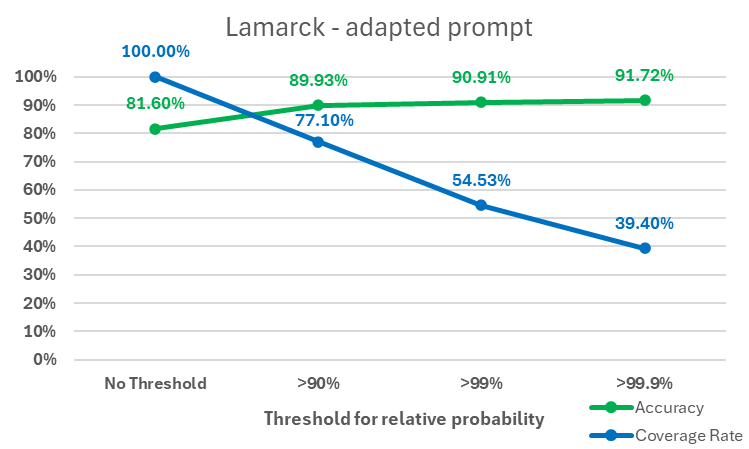

Supplementary Figure 13: Rate of accuracy and coverage for the classifications using the adapted prompt with the Lamarck model, depending on the threshold for the relative probability.
