## Supplementary Tables for "Implementing a Resource-Light and Low-Code Large Language Model System for Information Extraction from Mammography Reports: A Case Study"

Supplementary Table 1: Inter-Rater Agreement between the two researchers assigning the CDE values calculating Cohen’s kappa. *For some CDEs Cohen’s kappa cannot be calculated due to low occurrence in the analyzed data set with not all values appearing in the data

|  | | | **Cohen’s Kappa** |
| --- | --- | --- | --- |
| **Anamnesis** | | | **0.90** |
|  | **Family anamnesis** | | **0.94** |
|  |  | Positive family anamnesis mentioned | **0.95** |
|  |  | Relative with breast cancer | **0.89** |
|  | **Therapeutic anamnesis** | | **0.90** |
|  |  | Known breast cancer | **1.00** |
|  |  | Laterality of known breast cancer | **1.00** |
|  |  | Previous irradiation | **0.72** |
|  |  | Previous chemotherapy | **0.68** |
|  |  | Previous surgery | **0.88** |
|  |  | Second known breast cancer | **1.00** |
|  |  | Laterality of second known breast cancer | **-*** |
|  |  | Irradiation of second known breast cancer | **0.00** |
|  |  | Chemotherapy for second known breast cancer | **-*** |
|  |  | Surgery for second known breast cancer | **-*** |
|  |  | Non-oncological surgery conducted | **0.31** |
|  |  | Previous mammography conducted | **0.90** |
|  |  | Previous MR-mammography conducted | **0.73** |
|  |  | Previous biopsy conducted | **0.85** |
|  |  | Laterality of previously conducted biopsy | **1.00** |
|  |  | Quadrant of previously conducted biopsy | **0.00** |
|  |  | Clock position of previously conducted biopsy | **1.00** |
|  |  | Depth of previously conducted biopsy | **-*** |
|  |  | Result of previously conducted biopsy | **1.00** |
|  |  | Previous sonography conducted | **0.87** |
| **Report** | | |  |
|  | **Breast composition** | | **0.92** |
|  |  | ACR right breast | **0.91** |
|  |  | ACR left breast | **0.93** |
|  | **Findings** | | **0.71** |
|  |  | Lesion mentioned | **0.80** |
|  |  | Shape of the first lesion mentioned | **0.47** |
|  |  | Margin of the first lesion mentioned | **0.69** |
|  |  | Density of the first lesion mentioned | **0.43** |
|  |  | Dynamic of the first lesion mentioned | **0.76** |
|  |  | Mentioning of associated calcifications of the first lesion mentioned | **0.52** |
|  |  | Laterality of the first lesion mentioned | **0.92** |
|  |  | Visibility on view of the first lesion mentioned | **0.00** |
|  |  | Quadrant of the first lesion mentioned | **0.07** |
|  |  | Clock position of the first lesion mentioned | **0.72** |
|  |  | Depth of the first lesion mentioned | **0.80** |
|  |  | Second lesion mentioned | **0.90** |
|  |  | Shape of the second lesion mentioned | **0.60** |
|  |  | Margin of the second lesion mentioned | **0.59** |
|  |  | Density of the second lesion mentioned | **0.00** |
|  |  | Dynamic of the second lesion mentioned | **0.45** |
|  |  | Associated calcification mentioned of the second lesion mentioned | **0.75** |
|  |  | Laterality of the second lesion mentioned | **0.81** |
|  |  | Visibility on view of the second lesion mentioned | **0.00** |
|  |  | Quadrant of the second lesion mentioned | **0.37** |
|  |  | Clock position of the second lesion mentioned | **0.79** |
|  |  | Depth of the second lesion mentioned | **0.00** |
|  |  | Third lesion mentioned | **0.00** |
|  |  | Shape of the third lesion mentioned | **-*** |
|  |  | Margin of the third lesion mentioned | **-*** |
|  |  | Density of the third lesion mentioned | **-*** |
|  |  | Dynamic of the third lesion mentioned | **0.00** |
|  |  | Associated calcification of the third lesion mentioned | **0.00** |
|  |  | Laterality of the third lesion mentioned | **0.00** |
|  |  | Visibility on view of the third lesion mentioned | **-*** |
|  |  | Quadrant of the third lesion mentioned | **0.00** |
|  |  | Clock position of the third lesion mentioned | **0.00** |
|  |  | Depth of the third lesion mentioned | **-*** |
|  |  | Asymmetry mentioned | **0.91** |
|  |  | Description of mentioned asymmetry | **0.33** |
|  |  | Associated calcification of mentioned asymmetry | **1.00** |
|  |  | Laterality of mentioned asymmetry | **1.00** |
|  |  | Quadrant of mentioned asymmetry | **0.37** |
|  |  | Clock position of mentioned asymmetry | **1.00** |
|  |  | Depth of mentioned asymmetry | **0.00** |
|  |  | Architectural distorsion mentioned | **0.78** |
|  |  | Associated calcification of architectural distorsion mentioned | **0.67** |
|  |  | Depth of architectural distorsion | **0.00** |
|  |  | Laterality of architectural distorsion | **0.41** |
|  |  | Visibility on view of architectural distorsion | **0.00** |
|  |  | Quadrant of architectural distorsion | **0.00** |
|  |  | Clock position of architectural distorsion | **0.33** |
|  |  | Intramammary lymph node mentioned | **0.38** |
|  |  | Laterality of intramammary lymph node | **0.00** |
|  |  | Quadrant of intramammary lymph node | **0.00** |
|  |  | Clock position of intramammary lymph node | **-*** |
|  |  | Depth of intramammary lymph node | **0.00** |
|  |  | Solitary dilated duct | **-*** |
|  | **BIRADS** | | **0.95** |
|  |  | BIRADS left breast | **0.98** |
|  |  | BIRADS right breast | **0.92** |
| **Overall** | | | **0.83** |
