## Appendix 1 for "Implementing a Resource-Light and Low-Code Large Language Model System for Information Extraction from Mammography Reports: A Case Study"

**Appendix 1: CDEs used in the data structure**

**Group ‘Family anamnesis’**

- **Positive family anamnesis**
  - Type: Value List
  - Permissible Values: ‘yes’ (ja), ‘no’ (nein)
  - Description: Determines whether there is a mentioning of a blood-related family member in the report who already had a breast cancer disease.
- **Relative with breast cancer**
  - Type: Value List
  - Permissible Values: ‘mother’ (Mutter), ‘aunt’ (Tante), ‘grandmother’ (Grossmutter), ‘sister’ (Schwester), ‘other’ (andere), ‘unclear’ (unklar)
  - Description: Description, what is the relationship to the person with a breast cancer.
  - Condition: Positive family anamnesis = ‘yes’
  - Notes: Of course, it is possible that several relatives have breast cancer and not just one individual. As in our dataset there were no cases with more than one relative with breast cancer mentioned, this was not addressed as part of this study. In principle this could be done similar as it has been resolved for other factors with multiple occurrences (like multiple breast cancer diseases or multiple lesions; see below).

**Group ‘Therapeutic anamnesis’**

**Subgroup ‘Known breast cancer’**

- **Known breast cancer (1)**
  - Type: Value List
  - Permissible Values: ‘yes’ (ja), ‘no’ (nein)
  - Description: Determines whether there is already a known breast cancer mentioned.
- **Laterality of known breast cancer (1)**
  - Type: Value List
  - Permissible Values: ‘left’ (links), ‘right’ (rechts), ‘not mentioned’ (nicht erwaehnt)
  - Description: The laterality of the known breast cancer disease mentioned. In case there are multiple breast cancer diseases mentioned it is related to the first disease mentioned in the text.
  - Condition: Known breast cancer (1) = ‘yes’
- **Known breast cancer (2)**
  - Type: Value List
  - Permissible Values: ‘yes’ (ja), ‘no’ (nein)
  - Description: Determines whether there is a second known breast cancer mentioned (requiring that a first one is mentioned; second one in terms of appearance order in the text).
  - Condition: Known breast cancer (1) = ‘yes’
- **Laterality of known breast cancer (2)**
  - Type: Value List
  - Permissible Values: ‘left’ (links), ‘right’ (rechts), ‘not mentioned’ (nicht erwaehnt)
  - Description: The laterality of the second known breast cancer disease mentioned.
  - Condition: Known breast cancer (2) = ‘yes’

**Subgroup ‘Previous therapies’**

- **Previous radiotherapy (1)**
  - Type: Value List
  - Permissible Values: ‘yes’ (ja), ‘no’ (nein)
  - Description: Determines whether there is a mentioning of a previously conducted radiotherapy as therapy for a breast cancer in the report.
  - Condition: Known breast cancer (1) = ‘yes’
- **Previous chemotherapy (1)**
  - Type: Value List
  - Permissible Values: ‘yes’ (ja), ‘no’ (nein)
  - Description: Determines whether there is a mentioning of a previously conducted chemotherapy as therapy for a breast cancer in the report.
  - Condition: Known breast cancer (1) = ‘yes’
- **Previous oncological surgery (1)**
  - Type: Value List
  - Permissible Values: ‘yes’ (ja), ‘no’ (nein)
  - Description: Determines whether there is a mentioning of a previously conducted breast surgery as therapy for a breast cancer in the report.
  - Condition: Known breast cancer (1) = ‘yes’
- **Previous radiotherapy (2)**
  - Type: Value List
  - Permissible Values: ‘yes’ (ja), ‘no’ (nein)
  - Description: Determines whether there is a mentioning of a second previously conducted radiotherapy as therapy for a breast cancer in the report.
  - Condition: Known breast cancer (2) = ‘yes’
- **Previous chemotherapy (2)**
  - Type: Value List
  - Permissible Values: ‘yes’ (ja), ‘no’ (nein)
  - Description: Determines whether there is a mentioning of a second previously conducted chemotherapy as therapy for a breast cancer in the report.
  - Condition: Known breast cancer (2) = ‘yes’
- **Previous oncological surgery (2)**
  - Type: Value List
  - Permissible Values: ‘yes’ (ja), ‘no’ (nein)
  - Description: Determines whether there is a mentioning of a second previously conducted breast surgery as therapy for a breast cancer in the report.
  - Condition: Known breast cancer (2) = ‘yes’
- **Non oncological surgery conducted**
  - Type: Value List
  - Permissible Values: ‘yes’ (ja), ‘no’ (nein)
  - Description: Determines whether there is a mentioning of a non-oncological surgery in the report

**Subgroup ‘Previous interventions’**

- **Previous mammography**
  - Type: Value List
  - Permissible Values: ‘yes’ (ja), ‘no’ (nein)
  - Description: Determines whether there is a mentioning of a previously conducted mammography in the report.
- **Previous MR mammography**
  - Type: Value List
  - Permissible Values: ‘yes’ (ja), ‘no’ (nein)
  - Description: Determines whether there is a mentioning of a previously conducted MR mammography in the report.
- **Previous biopsy**
  - Type: Value List
  - Permissible Values: ‘yes’ (ja), ‘no’ (nein)
  - Description: Determines whether there is a mentioning of a previously conducted biopsy in the report.
- **Laterality of previous biopsy**
  - Type: Value List
  - Permissible Values: ‘left’ (links), ‘right’ (rechts), ‘not mentioned’ (nicht erwaehnt)
  - Description: Determines the laterality of the previous conducted biopsy.
  - Condition: Previous biopsy = ‘yes’
- **Quadrant of previous biopsy**
  - Type: Value List
  - Permissible Values: ‘upper outer’ (oben aussen), ‘upper inner’ (oben innen), ´lower outer‘ (unten aussen), ‘lower inner‘ (unten innen), ‘not mentioned’ (nicht erwaehnt)
  - Description: Determines the quadrant of the previous conducted biopsy.
  - Condition: Previous biopsy = ‘yes’
- **Clock position of previous biopsy**
  - Type: Value List
  - Permissible Values: ‘1’, ‘2’, ‘3’, ‘4’, ‘5’, ‘6’, ‘7’, ‘8’, ‘9’, ‘10’, ‘11’, ‘12’, ‘not mentioned’ (nicht erwaehnt)
  - Description: Determines the clock position of the previous conducted biopsy.
  - Condition: Previous biopsy = ‘yes’
- **Localization depth of the previous biopsy**
  - Type: Value List
  - Permissible Values: ‘Anterior’, ‘Middle’ (Mitte), ‘Posterior’, ‘Prepectoral’ (Präpektoral), ‘not mentioned’ (nicht erwaehnt)
  - Description: Determines the localization depth of the previous conducted biopsy.
  - Condition: Previous biopsy = ‘yes’
- **Result of previous biopsy**
  - Type: Value List
  - Permissible Values: ‘Breast cancer’ (Brustkrebs), ‘benign’ (Benigne), ‘unclear’ (nicht aussagekräftig), ‘not mentioned’ (nicht erwaehnt)
  - Description: Result of the previous biopsy.
  - Condition: Previous biopsy = ‘yes’
- **Previous sonography**
  - Type: Value List
  - Permissible Values: ‘yes’ (ja), ‘no’ (nein)
  - Description: Determines whether there is a mentioning of a previously conducted sonography in the report.

**Group ‘Breast composition’**

- **ACR category right breast**
  - Type: Value List
  - Permissible Values: ‘A’, ‘B’, ‘C’, ‘D’, ‘not mentioned’ (nicht erwaehnt)
  - Description: The mentioned ACR category of the right breast
- **ACR category left breast**
  - Type: Value List
  - Permissible Values: ‘A’, ‘B’, ‘C’, ‘D’, ‘not mentioned’ (nicht erwaehnt)
  - Description: The mentioned ACR category of the left breast

**Group ‘Findings’**

**Subgroup ‘Lesions’**

- **Lesion mentioned (1)**
  - Type: Value List
  - Permissible Values: ‘yes’ (ja), ‘no’ (nein)
  - Description: Determines whether there is a mentioning of a lesion in the report.
- **Lesion – shape (1)**
  - Type: Value List
  - Permissible Values: ‘oval’, ‘round’ (rund), ‘irregular’ (irregular), ‘not mentioned’ (nicht erwaehnt)
  - Description: Determines the shape of the mentioned lesion in the report.
  - Condition: Lesion mentioned (1) = ‘yes’
- **Lesion – margin (1)**
  - Type: Value List
  - Permissible Values: ‘circumscribed’ (umschrieben), ‘darkened’ (verdunkelt), ‘microlobulated’ (mikrolobuliert), ‘unclear’ (unklar), ‘spiculated’ (spikuliert), ‘not mentioned’ (nicht erwaehnt)
  - Description: The margin of the mentioned lesion in the report.
  - Condition: Lesion mentioned (1) = ‘yes’
- **Lesion – density (1)**
  - Type: Value List
  - Permissible Values: ‘hyperdense’, ‘isodens’, ‘hypodens’, ‘fatty’ (fetthaltig), ‘not mentioned’ (nicht erwaehnt)
  - Description: The density of the mentioned lesion in the report.
  - Condition: Lesion mentioned (1) = ‘yes’
- **Lesion – dynamic (1)**
  - Type: Value List
  - Permissible Values: ‘progredient’, ‘regredient’, ‘new’ (neu), ‘not mentioned’ (nicht erwaehnt)
  - Description: The dynamic of the mentioned lesion in the report.
  - Condition: Lesion mentioned (1) = ‘yes’
- **Lesion – associated calcification (1)**
  - Type: Value List
  - Permissible Values: ‘yes’ (ja), ‘no’ (nein), ‘not mentioned’ (nicht erwaehnt)
  - Description: Determines whether there are associated calcifications of the lesion mentioned.
  - Condition: Lesion mentioned (1) = ‘yes’
- **Lesion – laterality (1)**

Type: Value List

- Permissible Values: ‘left’ (links), ‘right’ (rechts), ‘not mentioned’ (nicht erwaehnt)
- Description: Determines the laterality of the lesion mentioned.
- Condition: Lesion mentioned (1) = ‘yes’
- **Lesion – visibility on view (1)**
  - Type: Value List
  - Permissible Values: ‘craniocaudal’ (kraniokaudal), ‘mediolateral oblique’ (mediolateral oblique), ‘not mentioned’ (nicht erwaehnt)
  - Description: The visibility of the mentioned lesion on the views in the mammography.
  - Condition: Lesion mentioned (1) = ‘yes’
- **Lesion – quadrant (1)**
  - Type: Value List
  - Permissible Values: ‘upper outer’ (oben aussen), ‘upper inner’ (oben innen), ´lower outer‘ (unten aussen), ‘lower inner‘ (unten innen), ‘not mentioned’ (nicht erwaehnt)
  - Description: Determines the quadrant of the mentioned lesion.
  - Condition: Lesion mentioned (1) = ‘yes’
- **Lesion - clock position (1)**
  - Type: Value List
  - Permissible Values: ‘1’, ‘2’, ‘3’, ‘4’, ‘5’, ‘6’, ‘7’, ‘8’, ‘9’, ‘10’, ‘11’, ‘12’, ‘not mentioned’ (nicht erwaehnt)
  - Description: Determines the clock position of the mentioned lesion.
  - Condition: Lesion mentioned (1) = ‘yes’
- **Lesion – depth of localization (1)**
  - Type: Value List
  - Permissible Values: ‘Anterior’, ‘Middle’ (Mitte), ‘Posterior’, ‘Prepectoral’ (Präpektoral), ‘not mentioned’ (nicht erwaehnt)
  - Description: Determines the localization depth of the mentioned lesion
  - Condition: Lesion mentioned (1) = ‘yes’
- **Lesion mentioned (2)**
  - Type: Value List
  - Permissible Values: ‘yes’ (ja), ‘no’ (nein)
  - Description: Determines whether there is a mentioning of a second lesion in the report (order of appearance in the report).
- **Lesion – shape (2)**
  - Type: Value List
  - Permissible Values: ‘oval’, ‘round’ (rund), ‘irregular’ (irregular), ‘not mentioned’ (nicht erwaehnt)
  - Description: Determines the shape of the second lesion mentioned in the report.
  - Condition: Lesion mentioned (2) = ‘yes’
- **Lesion – margin (2)**
  - Type: Value List
  - Permissible Values: ‘circumscribed’ (umschrieben), ‘darkened’ (verdunkelt), ‘microlobulated’ (mikrolobuliert), ‘unclear’ (unklar), ‘spiculated’ (spikuliert), ‘not mentioned’ (nicht erwaehnt)
  - Description: The margin of the second lesion mentioned in the report.
  - Condition: Lesion mentioned (2) = ‘yes’
- **Lesion – density (2)**
  - Type: Value List
  - Permissible Values: ‘hyperdense’, ‘isodens’, ‘hypodens’, ‘fatty’ (fetthaltig), ‘not mentioned’ (nicht erwaehnt)
  - Description: The density of the second lesion mentioned in the report.
  - Condition: Lesion mentioned (2) = ‘yes’
- **Lesion – dynamic (2)**
  - Type: Value List
  - Permissible Values: ‘progredient’, ‘regredient’, ‘new’ (neu), ‘not mentioned’ (nicht erwaehnt)
  - Description: The dynamic of the second lesion mentioned in the report.
  - Condition: Lesion mentioned (2) = ‘yes’
- **Lesion – associated calcification (2)**
  - Type: Value List
  - Permissible Values: ‘yes’ (ja), ‘no’ (nein), ‘not mentioned’ (nicht erwaehnt)
  - Description: Determines whether there are associated calcifications of the second lesion mentioned in the report.
  - Condition: Lesion mentioned (2) = ‘yes’
- **Lesion – laterality (2)**

Type: Value List

- Permissible Values: ‘left’ (links), ‘right’ (rechts), ‘not mentioned’ (nicht erwaehnt)
- Description: Determines the laterality of the second lesion mentioned in the report.
- Condition: Lesion mentioned (2) = ‘yes’
- **Lesion – visibility on view (2)**
  - Type: Value List
  - Permissible Values: ‘craniocaudal’ (kraniokaudal), ‘mediolateral oblique’ (mediolateral oblique), ‘not mentioned’ (nicht erwaehnt)
  - Description: The visibility of the second lesion mentioned on the views in the mammography.
  - Condition: Lesion mentioned (2) = ‘yes’
- **Lesion – quadrant (2)**
  - Type: Value List
  - Permissible Values: ‘upper outer’ (oben aussen), ‘upper inner’ (oben innen), ´lower outer‘ (unten aussen), ‘lower inner‘ (unten innen), ‘not mentioned’ (nicht erwaehnt)
  - Description: Determines the quadrant of the second lesion mentioned.
  - Condition: Lesion mentioned (2) = ‘yes’
- **Lesion - clock position (2)**
  - Type: Value List
  - Permissible Values: ‘1’, ‘2’, ‘3’, ‘4’, ‘5’, ‘6’, ‘7’, ‘8’, ‘9’, ‘10’, ‘11’, ‘12’, ‘not mentioned’ (nicht erwaehnt)
  - Description: Determines the clock position of the second lesion mentioned.
  - Condition: Lesion mentioned (2) = ‘yes’
- **Lesion – depth of localization (2)**
  - Type: Value List
  - Permissible Values: ‘Anterior’, ‘Middle’ (Mitte), ‘Posterior’, ‘Prepectoral’ (Präpektoral), ‘not mentioned’ (nicht erwaehnt)
  - Description: Determines the localization depth of the second lesion mentioned.
  - Condition: Lesion mentioned (2) = ‘yes’
- **Lesion mentioned (3)**
  - Type: Value List
  - Permissible Values: ‘yes’ (ja), ‘no’ (nein)
  - Description: Determines whether there is a mentioning of a third lesion in the report (order of appearance in the report).
- **Lesion – shape (3)**
  - Type: Value List
  - Permissible Values: ‘oval’, ‘round’ (rund), ‘irregular’ (irregular), ‘not mentioned’ (nicht erwaehnt)
  - Description: Determines the shape of the third lesion mentioned in the report.
  - Condition: Lesion mentioned (3) = ‘yes’
- **Lesion – margin (3)**
  - Type: Value List
  - Permissible Values: ‘circumscribed’ (umschrieben), ‘darkened’ (verdunkelt), ‘microlobulated’ (mikrolobuliert), ‘unclear’ (unklar), ‘spiculated’ (spikuliert), ‘not mentioned’ (nicht erwaehnt)
  - Description: The margin of the third lesion mentioned in the report.
  - Condition: Lesion mentioned (3) = ‘yes’
- **Lesion – density (3)**
  - Type: Value List
  - Permissible Values: ‘hyperdense’, ‘isodens’, ‘hypodens’, ‘fatty’ (fetthaltig), ‘not mentioned’ (nicht erwaehnt)
  - Description: The density of the third lesion mentioned in the report.
  - Condition: Lesion mentioned (3) = ‘yes’
- **Lesion – dynamic (3)**
  - Type: Value List
  - Permissible Values: ‘progredient’, ‘regredient’, ‘new’ (neu), ‘not mentioned’ (nicht erwaehnt)
  - Description: The dynamic of the third lesion mentioned in the report.
  - Condition: Lesion mentioned (3) = ‘yes’
- **Lesion – associated calcification (3)**
  - Type: Value List
  - Permissible Values: ‘yes’ (ja), ‘no’ (nein), ‘not mentioned’ (nicht erwaehnt)
  - Description: Determines whether there are associated calcifications of the third lesion mentioned in the report.
  - Condition: Lesion mentioned (3) = ‘yes’
- **Lesion – laterality (3)**

Type: Value List

- Permissible Values: ‘left’ (links), ‘right’ (rechts), ‘not mentioned’ (nicht erwaehnt)
- Description: Determines the laterality of the third lesion mentioned in the report.
- Condition: Lesion mentioned (3) = ‘yes’
- **Lesion – visibility on view (3)**
  - Type: Value List
  - Permissible Values: ‘craniocaudal’ (kraniokaudal), ‘mediolateral oblique’ (mediolateral oblique), ‘not mentioned’ (nicht erwaehnt)
  - Description: The visibility of the third lesion mentioned on the views in the mammography.
  - Condition: Lesion mentioned (3) = ‘yes’
- **Lesion – quadrant (3)**
  - Type: Value List
  - Permissible Values: ‘upper outer’ (oben aussen), ‘upper inner’ (oben innen), ´lower outer‘ (unten aussen), ‘lower inner‘ (unten innen), ‘not mentioned’ (nicht erwaehnt)
  - Description: Determines the quadrant of the third lesion mentioned.
  - Condition: Lesion mentioned (3) = ‘yes’
- **Lesion - clock position (3)**
  - Type: Value List
  - Permissible Values: ‘1’, ‘2’, ‘3’, ‘4’, ‘5’, ‘6’, ‘7’, ‘8’, ‘9’, ‘10’, ‘11’, ‘12’, ‘not mentioned’ (nicht erwaehnt)
  - Description: Determines the clock position of the third lesion mentioned.
  - Condition: Lesion mentioned (3) = ‘yes’
- **Lesion – depth of localization (3)**
  - Type: Value List
  - Permissible Values: ‘Anterior’, ‘Middle’ (Mitte), ‘Posterior’, ‘Prepectoral’ (Präpektoral), ‘not mentioned’ (nicht erwaehnt)
  - Description: Determines the localization depth of the third lesion mentioned.
  - Condition: Lesion mentioned (3) = ‘yes’

**Subgroup ‘Asymmetry’**

- **Asymmetry mentioned**
  - Type: Value List
  - Permissible Values: ‘yes’ (ja), ‘no’ (nein)
  - Description: Determines whether there is a mentioning of an asymmetry in the report.
- **Asymmetry – shape**
  - Type: Value List
  - Permissible Values: ‘oval’, ‘round’ (rund), ‘irregular’ (irregular), ‘not mentioned’ (nicht erwaehnt)
  - Description: Determines the shape of the mentioned asymmetry in the report.
  - Condition: Asymmetry mentioned = ‘yes’
- **Asymmetry – associated calcification**
  - Type: Value List
  - Permissible Values: ‘yes’ (ja), ‘no’ (nein), ‘not mentioned’ (nicht erwaehnt)
  - Description: Determines whether there are associated calcifications of the asymmetry mentioned.
  - Condition: Asymmetry mentioned = ‘yes’
- **Asymmetry – laterality**

Type: Value List

- Permissible Values: ‘left’ (links), ‘right’ (rechts), ‘not mentioned’ (nicht erwaehnt)
- Description: Determines the laterality of the asymmetry mentioned.
- Condition: Asymmetry mentioned = ‘yes’
- **Asymmetry – quadrant**
  - Type: Value List
  - Permissible Values: ‘upper outer’ (oben aussen), ‘upper inner’ (oben innen), ´lower outer‘ (unten aussen), ‘lower inner‘ (unten innen), ‘not mentioned’ (nicht erwaehnt)
  - Description: Determines the quadrant of the mentioned asymmetry.
  - Condition: Asymmetry mentioned = ‘yes’
- **Asymmetry - clock position**
  - Type: Value List
  - Permissible Values: ‘1’, ‘2’, ‘3’, ‘4’, ‘5’, ‘6’, ‘7’, ‘8’, ‘9’, ‘10’, ‘11’, ‘12’, ‘not mentioned’ (nicht erwaehnt)
  - Description: Determines the clock position of the mentioned asymmetry.
  - Condition: Asymmetry mentioned = ‘yes’
- **Asymmetry – depth of localization**
  - Type: Value List
  - Permissible Values: ‘Anterior’, ‘Middle’ (Mitte), ‘Posterior’, ‘Prepectoral’ (Präpektoral), ‘not mentioned’ (nicht erwaehnt)
  - Description: Determines the localization depth of the mentioned asymmetry.
  - Condition: Asymmetry mentioned = ‘yes’

**Subgroup ‘Architectural distorsion’**

- **Architectural distorsion mentioned**
  - Type: Value List
  - Permissible Values: ‘yes’ (ja), ‘no’ (nein)
  - Description: Determines whether there is a mentioning of an architectural distorsion in the report.
- **Architectural distorsion – associated calcification**
  - Type: Value List
  - Permissible Values: ‘yes’ (ja), ‘no’ (nein), ‘not mentioned’ (nicht erwaehnt)
  - Description: Determines whether there are associated calcifications of the architectural distorsion mentioned.
  - Condition: Architectural distorsion mentioned = ‘yes’
- **Architectural distorsion – laterality**

Type: Value List

- Permissible Values: ‘left’ (links), ‘right’ (rechts), ‘not mentioned’ (nicht erwaehnt)
- Description: Determines the laterality of the architectural distorsion mentioned.
- Condition: Architectural distorsion mentioned = ‘yes’
- **Architectural distorsion – quadrant**
  - Type: Value List
  - Permissible Values: ‘upper outer’ (oben aussen), ‘upper inner’ (oben innen), ´lower outer‘ (unten aussen), ‘lower inner‘ (unten innen), ‘not mentioned’ (nicht erwaehnt)
  - Description: Determines the quadrant of the mentioned architectural distorsion.
  - Condition: Architectural distorsion mentioned = ‘yes’
- **Architectural distorsion - clock position**
  - Type: Value List
  - Permissible Values: ‘1’, ‘2’, ‘3’, ‘4’, ‘5’, ‘6’, ‘7’, ‘8’, ‘9’, ‘10’, ‘11’, ‘12’, ‘not mentioned’ (nicht erwaehnt)
  - Description: Determines the clock position of the mentioned Architectural distorsion.
  - Condition: Architectural distorsion mentioned = ‘yes’
- **Architectural distorsion – depth of localization**
  - Type: Value List
  - Permissible Values: ‘Anterior’, ‘Middle’ (Mitte), ‘Posterior’, ‘Prepectoral’ (Präpektoral), ‘not mentioned’ (nicht erwaehnt)
  - Description: Determines the localization depth of the mentioned architectural distorsion.
  - Condition: Architectural distorsion mentioned = ‘yes’

**Subgroup ‘Intramammary lymph node’**

- **Intramammary lymph node mentioned**
  - Type: Value List
  - Permissible Values: ‘yes’ (ja), ‘no’ (nein)
  - Description: Determines whether there is a mentioning of an Intramammary lymph node in the report.
- **Intramammary lymph node – laterality**

Type: Value List

- Permissible Values: ‘left’ (links), ‘right’ (rechts), ‘not mentioned’ (nicht erwaehnt)
- Description: Determines the laterality of the Intramammary lymph node mentioned.
- Condition: Intramammary lymph node mentioned = ‘yes’
- **Intramammary lymph node – quadrant**
  - Type: Value List
  - Permissible Values: ‘upper outer’ (oben aussen), ‘upper inner’ (oben innen), ´lower outer‘ (unten aussen), ‘lower inner‘ (unten innen), ‘not mentioned’ (nicht erwaehnt)
  - Description: Determines the quadrant of the mentioned Intramammary lymph node.
  - Condition: Intramammary lymph node mentioned = ‘yes’
- **Intramammary lymph node - clock position**
  - Type: Value List
  - Permissible Values: ‘1’, ‘2’, ‘3’, ‘4’, ‘5’, ‘6’, ‘7’, ‘8’, ‘9’, ‘10’, ‘11’, ‘12’, ‘not mentioned’ (nicht erwaehnt)
  - Description: Determines the clock position of the mentioned Intramammary lymph node.
  - Condition: Intramammary lymph node mentioned = ‘yes’
- **Intramammary lymph node – depth of localization**
  - Type: Value List
  - Permissible Values: ‘Anterior’, ‘Middle’ (Mitte), ‘Posterior’, ‘Prepectoral’ (Präpektoral), ‘not mentioned’ (nicht erwaehnt)
  - Description: Determines the localization depth of the mentioned Intramammary lymph node.
  - Condition: Intramammary lymph node mentioned = ‘yes’

**Subgroup ‘Solitary dilated duct’**

- **Solitary dilated duct mentioned**
  - Type: Value List
  - Permissible Values: ‘yes’ (ja), ‘no’ (nein)
  - Description: Determines whether there is a mentioning of a solitary dilated duct in the report.

**Group ‘BIRADS’**

- **BIRADS left breast**
  - Type: Value List
  - Permissible Values: ‘0’, ‘1’, ‘2’, ‘3’, ‘4’, ‘4A’, ‘4B’, ‘4C’, ‘5’, ‘6’
  - Description: The mentioned BIRADS score of the left breast
- **BIRADS right breast**
  - Type: Value List
  - Permissible Values: ‘0’, ‘1’, ‘2’, ‘3’, ‘4’, ‘4A’, ‘4B’, ‘4C’, ‘5’, ‘6’
  - Description: The mentioned BIRADS score of the right breast
